# An evaluation of the adequacy of Indian national and state Essential Medicines Lists (EMLs) for palliative care medical needs - a comparative analysis

**DOI:** 10.1101/2024.08.26.24312600

**Authors:** Disha Agrawal, Divya Shrinivas, Parth Sharma, M R Rajagopal, Arun Ghoshal, Siddhesh Zadey

## Abstract

**Objectives:** Essential Medicines Lists (EMLs) guide the public sector procurement and supply of medications to impact access to adequate and appropriate palliative care drugs. This study evaluates the adequacy of India’s national and sub-national EMLs that can directly impact palliative care for 5.4 million patients.

**Methods:** In this qualitative document review, we compared Indian national, and state EMLs acquired from official government websites with the International Association for Hospice & Palliative Care (IAHPC) EML recommendations. We analysed data on the indication and formulation of drugs under the different categories of formulations present (all, some, and no), and drugs absent. Literature review and inputs from palliative care experts provided alternatives of absent medications to assess the adequacy of lists in managing the symptoms listed by IAPHC.

**Results:** We analysed 3 national and 25 state lists for 33 recommended drugs. The Central Government Health Services list had the maximum availability of all formulations of drugs (16 [48%]) nationally. Among states and union territories, the Delhi EML was the closest to IAHPC with 17 (52%) drugs with all formulations present. Nagaland had the most incomplete EML with only 3 (9%) drugs with all formulations present. No EML had all the recommended formulations of morphine. In one national and sixteen state EMLs, oral morphine was absent.

**Conclusion:** While Indian EMLs lack drugs for palliative care when compared with the IAHPC EML, symptom management is adequate. There is a need for countries with limited resources to modify the IAPHC list for their settings.

**What is already known on this topic:** Essential Medicines Lists (EMLs) are instrumental in guiding public sector procurement of drugs. The implementation of EMLs is known to improve drug availability and prescription practices. The rising burden of people requiring end-of-life care globally necessitates the availability of appropriate drugs for the medical management of symptoms, which can be achieved through their inclusion in local EMLs.

**What this study adds:** The national and sub-national EMLs of India do not fully adhere to the International Association for Hospice and Palliative Care (IAHPC) recommendations. However, they contain adequate drugs for the management of the listed symptoms. Additionally, the inclusion of various formulations of morphine remains a challenge to be addressed.

**How this study might affect research, practice or policy:** This study highlights the need to develop a fit-for-purpose EML for palliative care, taking into account the geographical variations in palliative care needs, and resource constraints in healthcare delivery at the state and country level.

## 1. INTRODUCTION

Palliative care deals with reducing serious health-related suffering through the early identification, and treatment of physical, psychosocial, or spiritual problems related to acute or chronic diseases. (1) Adequate access to and provision of palliative care improves quality of life (2), enables informed treatment-decision making (3), and reduces hospital readmissions and healthcare costs. (4) Although access to palliative care is a right to health, globally, only 14% of the 40 million people in need can access it. (1)

Although 5.4 million Indians need palliative care annually, merely 1% can access it. (5) In Indian patients with end-stage cancers, the unmet need for palliative care was reported to be 98.3%. (6) Several obstacles disrupt effective palliative care delivery including poor geographical access, limited awareness, lack of workforce training, restrictive prescription policies for pain medications, and limited policy prioritisation, among others. (5)

The World Health Organization (WHO) introduced the concept of essential medicines in 1977 to address priority health needs. (7) These medications are chosen based on their public health importance, efficacy, safety, and cost-effectiveness, and should be consistently available in sufficient quantities in public health centres. Thus, the implementation of a thoughtfully curated Essential Medicines List (EML) can enhance the quality of care, management practices, and resource allocation, and ensure the availability of medicines by streamlining procurement and distribution processes. While countries can determine their EMLs, the WHO model list serves as a reference for national and institutional lists. (8)

India has multiple EMLs. Health is a state subject as per the Indian Constitution. Hence, states draft their EMLs to match the local needs. (9) Two national health insurance schemes - the Employees’ State Insurance Scheme (ESIS) and the Central Government Health Scheme (CGHS) have their EMLs. A third national EML exists for the states without their unique EML to follow. These EMLs guide the procurement of drugs that are dispensed at government-run healthcare institutions at the state and national levels. As the vast majority of Indians reside in rural areas and rely on public (government-run) healthcare facilities, the appropriateness of palliative care service delivery partly depends on the adequacy of the national and state EMLs.

In 2007, responding to a request from the World Health Organization’s (WHO) Cancer Control Program, the International Association for Hospice and Palliative Care (IAHPC) collaborated with other organisations to develop a list of essential medicines for the 16 most common symptoms of palliative care. (10) The IAHPC is a public charity, which serves as an international platform to improve access to palliative care and improve global standards of care. (11) This list thus became the model list to serve as a reference for nations globally.

The objective of this study was to evaluate the adequacy of India’s EMLs at the national and state/Union Territory (UT) levels by comparing Indian lists with the list curated by IAHPC. By identifying alternatives to missing drugs in the national and state lists, we also aimed to assess the adequacy of the lists for the management of common palliative care symptoms listed by IAHPC.

## 2. METHODS

### 2.1 Data sources

The National List of Essential Medicines (NLEM), last updated in 2022, was designed by the Ministry of Health and Family Welfare, Government of India. (12) The Employees’ State Insurance Scheme (ESIS) EML caters to the population employed in factories, and other establishments such as hotels, shops, and restaurants. (13) The Central Government Health Scheme (CGHS) covers current employees and pensioners of the national (central) government.

1. (14) The state/UT lists were designed for the respective state-level public health facilities.

The IAHPC list of essential medicines was drafted through the consensus of international physicians and pharmacologists. After identifying the most common symptoms in palliative care, a final list of appropriate medications was devised, using a modified Delphi process. (15) The IAHPC list included 33 essential drugs, which were looked for in individual national and state EMLs. We accessed the most recent versions of three national and 25 state/UT EMLs from the government websites **(Table 1)**. The links to access the individual EMLs are provided in **Supplementary Table 1**.

**Table 1:**
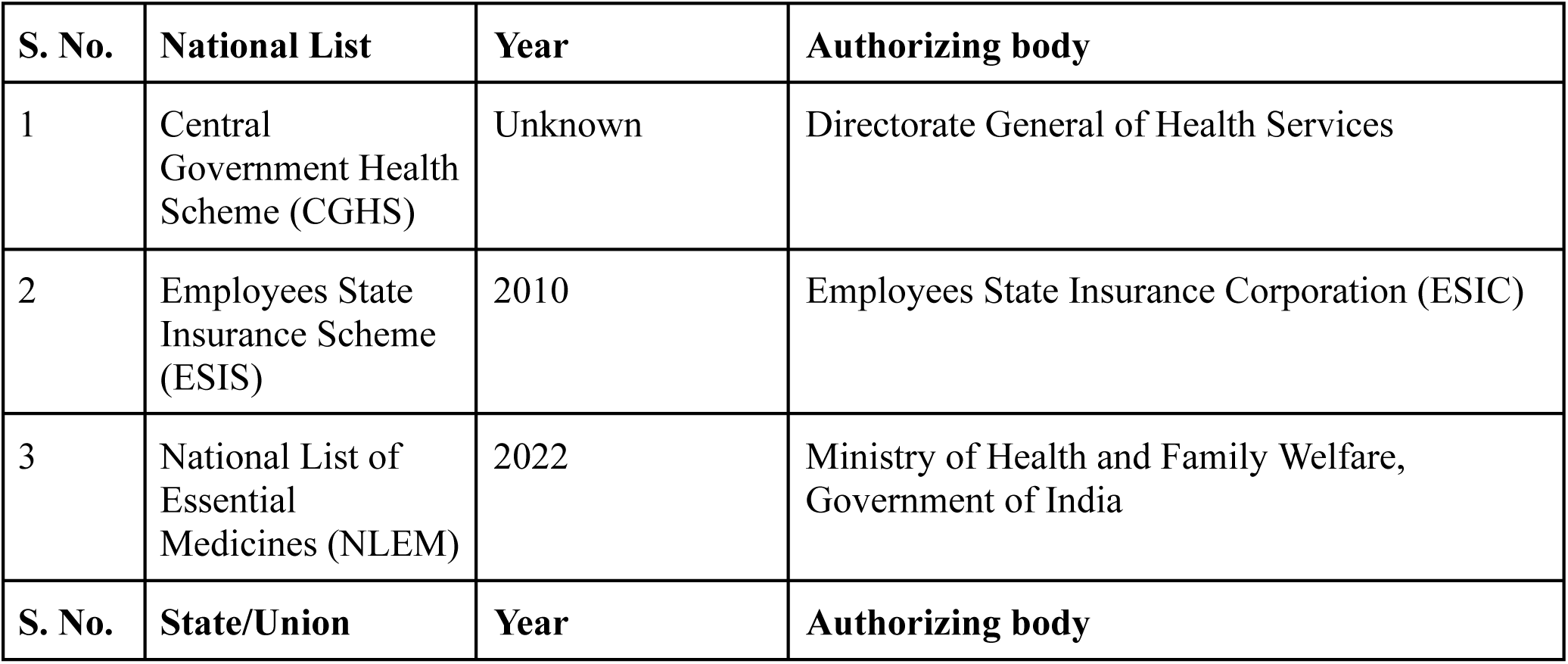
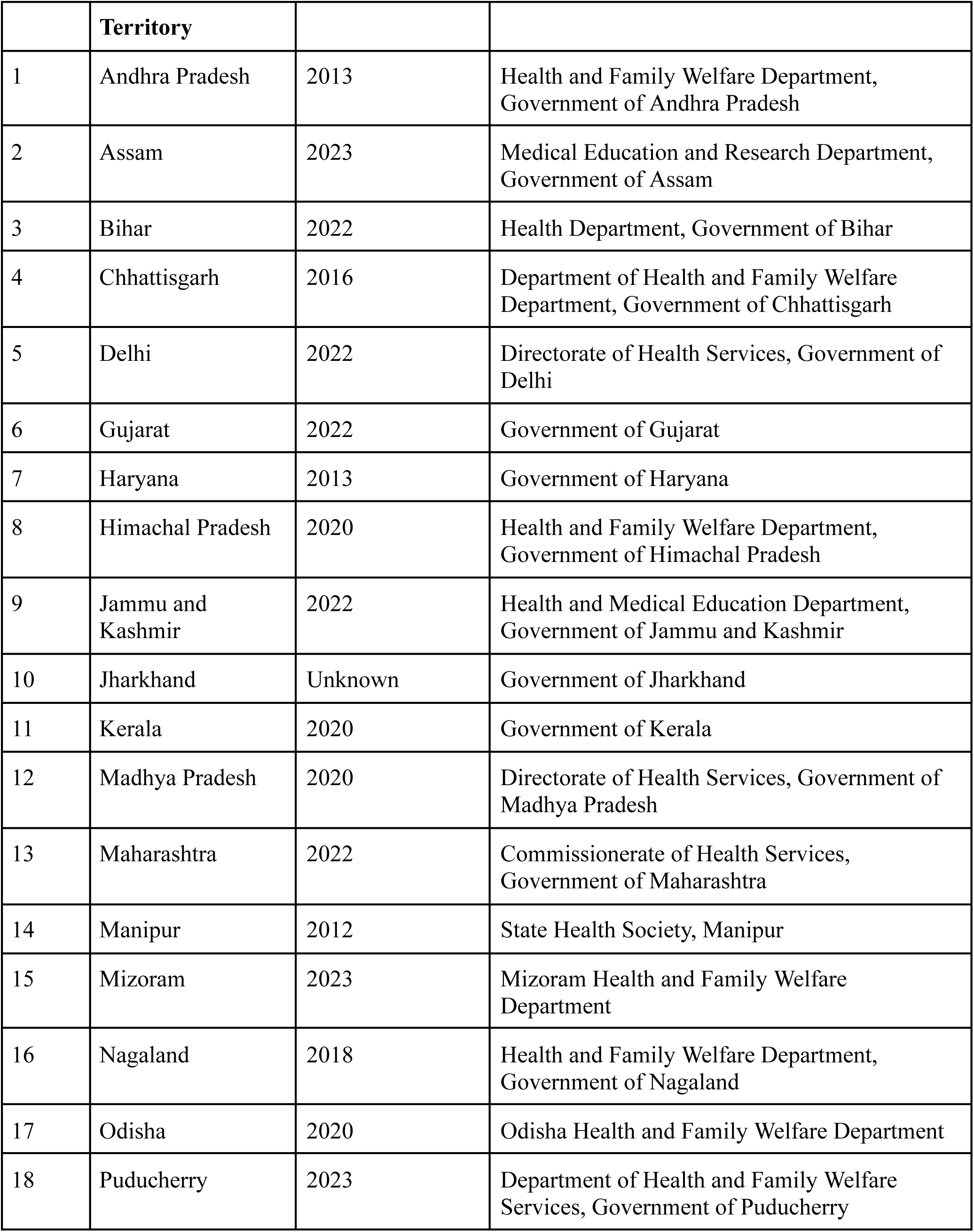
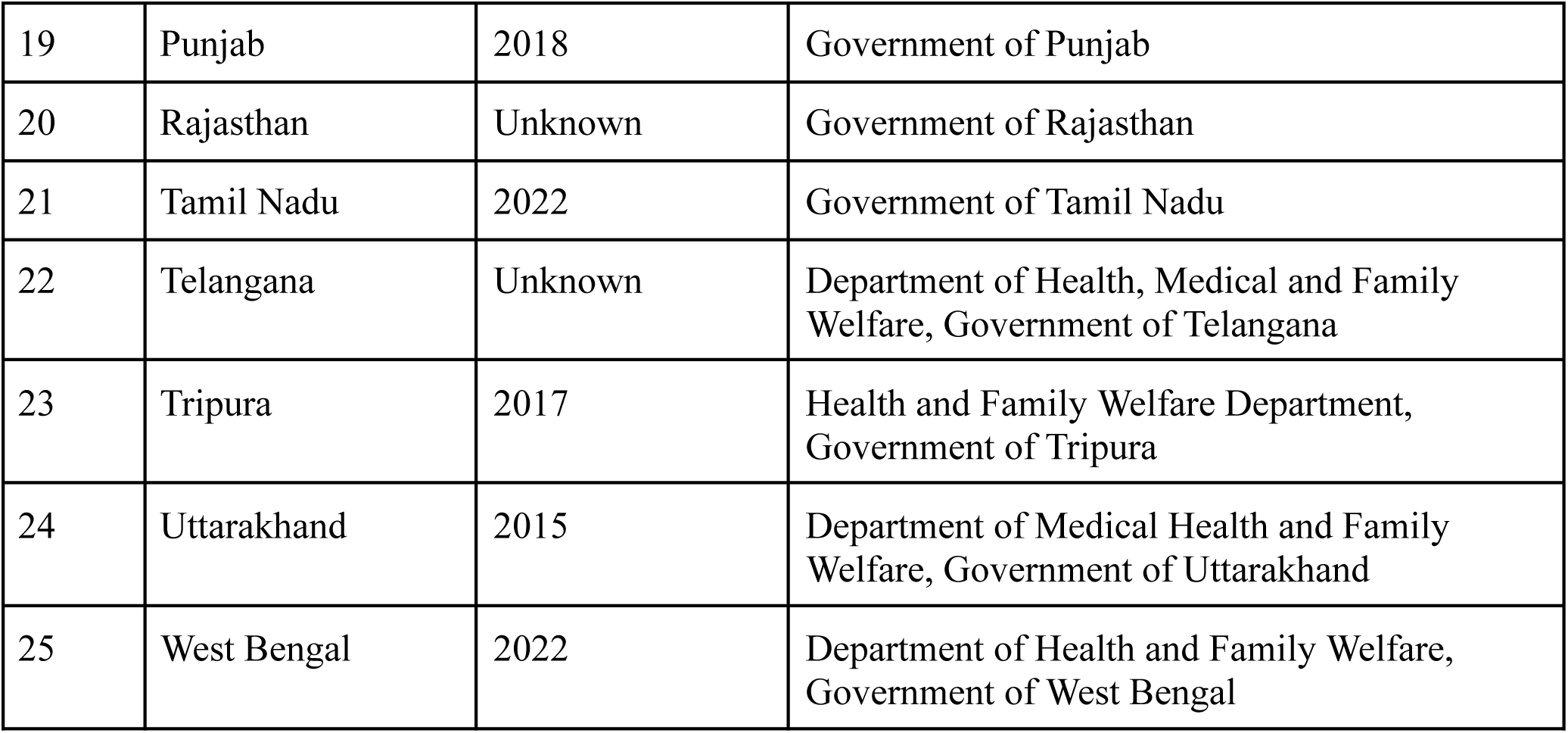
National and State Essential Medication Lists.

| S. No. | National List | Year | Authorizing body |
| --- | --- | --- | --- |
| 1 | Central Government Health Scheme (CGHS) | Unknown | Directorate General of Health Services |
| 2 | Employees State Insurance Scheme (ESIS) | 2010 | Employees State Insurance Corporation (ESIC) |
| 3 | National List of Essential Medicines (NLEM) | 2022 | Ministry of Health and Family Welfare, Government of India |
| S. No. | State/Union | Year | Authorizing body |

|  | **Territory** |  |  |
| --- | --- | --- | --- |
| 1 | Andhra Pradesh | 2013 | Health and Family Welfare Department,<br>Government of Andhra Pradesh |
| 2 | Assam | 2023 | Medical Education and Research Department,<br>Government of Assam |
| 3 | Bihar | 2022 | Health Department, Government of Bihar |
| 4 | Chhattisgarh | 2016 | Department of Health and Family Welfare<br>Department, Government of Chhattisgarh |
| 5 | Delhi | 2022 | Directorate of Health Services, Government of<br>Delhi |
| 6 | Gujarat | 2022 | Government of Gujarat |
| 7 | Haryana | 2013 | Government of Haryana |
| 8 | Himachal Pradesh | 2020 | Health and Family Welfare Department,<br>Government of Himachal Pradesh |
| 9 | Jammu and<br>Kashmir | 2022 | Health and Medical Education Department,<br>Government of Jammu and Kashmir |
| 10 | Jharkhand | Unknown | Government of Jharkhand |
| 11 | Kerala | 2020 | Government of Kerala |
| 12 | Madhya Pradesh | 2020 | Directorate of Health Services, Government of<br>Madhya Pradesh |
| 13 | Maharashtra | 2022 | Commissionerate of Health Services,<br>Government of Maharashtra |
| 14 | Manipur | 2012 | State Health Society, Manipur |
| 15 | Mizoram | 2023 | Mizoram Health and Family Welfare<br>Department |
| 16 | Nagaland | 2018 | Health and Family Welfare Department,<br>Government of Nagaland |
| 17 | Odisha | 2020 | Odisha Health and Family Welfare Department |
| 18 | Puducherry | 2023 | Department of Health and Family Welfare<br>Services, Government of Puducherry |
| 19 | Punjab | 2018 | Government of Punjab |
| 20 | Rajasthan | Unknown | Government of Rajasthan |
| 21 | Tamil Nadu | 2022 | Government of Tamil Nadu |
| 22 | Telangana | Unknown | Department of Health, Medical and Family Welfare, Government of Telangana |
| 23 | Tripura | 2017 | Health and Family Welfare Department, Government of Tripura |
| 24 | Uttarakhand | 2015 | Department of Medical Health and Family Welfare, Government of Uttarakhand |
| 25 | West Bengal | 2022 | Department of Health and Family Welfare, Government of West Bengal |

### 2.2 Data extraction

We followed the READ (readying material, extracting data, analysing data, and distilling findings) approach to evaluate national and state (including union territories) EMLs and compared them with the IAHPC EML of essential medicines for palliative care. (16) The drugs’ names and formulations present in the Indian EMLs were compiled in a Microsoft Excel spreadsheet and matched with the IAHPC list to assess the adequacy of the Indian EMLs.

If the EML did not have the drug, we looked for alternative drugs that may be prescribed for the same indication **(Table 2)**. Acceptable alternatives were determined by reviewing the literature and by consulting two experienced palliative care experts (MRR with 31 years of experience and AG with 12 years of experience). Recommendations for alternatives to any drug included in the IAHPC EML, but not present in an Indian EML, were taken from both experts individually in the first stage. Subsequently, both experts were invited to reach a consensus on any differences in recommendations in the second stage. The experts decided on alternative drugs after considering drug efficacy, safety, cost-effectiveness, availability in the Indian market, and secondary effects of the drug that would be useful in a patient receiving palliative care.

**Table 2:** Drugs identified for specific symptoms.

| S. No. | Symptom | Drugs in IAHPCL EML | Alternative drugs identified |
| --- | --- | --- | --- |
| 1 | Depression | Amitriptyline, | None |
|  |  | Citalopram (or any other equivalent generic SSRI except paroxetine and fluvoxamine),<br>Mirtazapine (or any other generic, dual action Nassa or SNRI) |  |
| 2 | Neuropathic pain | Amitriptyline,<br>Carbamazepine,<br>Dexamethasone,<br>Gabapentin | Pregabalin |
| 3 | Constipation | Bisacodyl, Senna | None |
| 4 | Diarrhoea | Codeine, Loperamide,<br>Octreotide | Diphenoxylate |
|  |  | Oral rehydration salts | None |
| 5 | Pain - mild to moderate | Codeine, Diclofenac,<br>Ibuprofen, Paracetamol,<br>Tramadol | Naproxen |
| 6 | Anorexia | Dexamethasone, Megestrol acetate, Prednisolone | None |
| 7 | Nausea | Dexamethasone,<br>Metoclopramide,<br>Diphenhydramine,<br>Haloperidol, Hyoscine butylbromide | Ondansetron, Domperidone,<br>Promethazine, Olanzapine |
| 8 | Vomiting | Dexamethasone,<br>Metoclopramide,<br>Diphenhydramine,<br>Haloperidol, Hyoscine butylbromide, Octreotide | Ondansetron, Domperidone,<br>Promethazine, Olanzapine |
| 9 | Anxiety | Diazepam, Lorazepam,<br>Midazolam | None |
| 10 | Pain - moderate to severe | Morphine, Methadone,<br>Fentanyl (transdermal patch),<br>Oxycodone | None |
| 11 | Delirium | Haloperidol,<br>Levomepromazine | None |
| 12 | Terminal restlessness | Haloperidol,<br>Levomepromazine,<br>Midazolam | None |
| 13 | Terminal respiratory congestion | Hyoscine butylbromide | Glycopyrrolate |
| 14 | Visceral pain | Hyoscine butylbromide | Dicyclomine (only for colicky or spasmodic pain) |
| 15 | Insomnia | Lorazepam, Trazodone,<br>Zolpidem | Alprazolam, Zaleplon,<br>Eszopiclone |
| 16 | Dyspnea | Morphine | None |

### 2.4 Data analysis

To assess the adequacy of the lists, drugs present in the national and state EMLs were categorised as follows: all formulations present, some formulations present, no recommended formulation present, and drug absent. We calculated the percentage of drugs present in each category in national and state EMLs using **Equation 1**. The different formulations that were mentioned for drugs included tablets, capsules, oral solutions, injectables, suppositories, and salts.

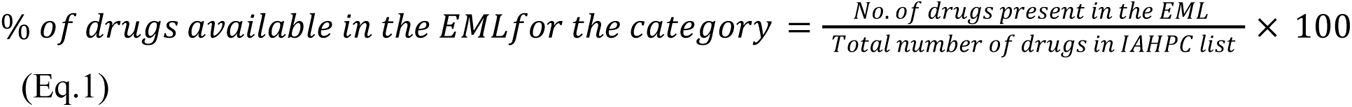

We also looked at the adequacy of the EMLs to manage palliative care symptoms using alternative drugs identified using expert consensus and literature review as mentioned previously. The IAHPC EML mentions 16 common palliative care symptoms which include depression, neuropathic pain, constipation, anorexia, nausea, vomiting, anxiety, mild to moderate pain, moderate to severe pain, delirium, terminal restlessness, terminal respiratory congestion, visceral pain, dyspnea, diarrhoea, and insomnia. We assessed whether the national or state EMLs had at least one drug that could be prescribed for each of these indications. For diarrhoea, the presence of ORS in the list was considered necessary to adequately manage the symptoms. For terminal restlessness, we considered haloperidol essential for management, with midazolam as an add-on drug. (17) We reported this result as the proportion of EMLs that were adequate to manage all the aforementioned symptoms.

The most recent National Programme for Prevention and Control of Non-Communicable Diseases (NPNCD) guidelines evaluate access to palliative care by assessing morphine-equivalent consumption of strong opioid analgesics (except methadone) per cancer death. (18) Therefore, we specifically looked at the inclusion of various formulations of morphine (oral solution, oral tablet, and injectable) in the EMLs as well. Subsequently, we calculated the proportion of EMLs which included both oral and injectable morphine, only oral morphine, only injectable morphine, and no formulation of morphine.

## 3. RESULTS

A total of 3 national and 25 state/UT EMLs were analysed in the study. Although India is a union of 28 states and 8 UTs, the remaining EMLs were not available in the public domain.

### 3.1 Adequacy of Drugs

Among national EMLs, the CGHS had the highest number of all formulations of drugs (16 [48%]). Ten (30%) drugs had some formulations present, one drug had no recommended formulation present, and six (19%) drugs were absent. The NLEM contained all formulations of 15 (46%) drugs, some formulations of 5 (15%) drugs, no recommended formulations of 1 (3%) drug and 12 (36%) drugs were absent. The ESIS EML had the least number of drugs with all formulations present (6 [18%]), some formulations were present of 7 (21%) drugs, no recommended formulations were present of 2 (6%) drugs, and 18 (55%) drugs were absent. **(Figure 1)**

**Figure 1:**
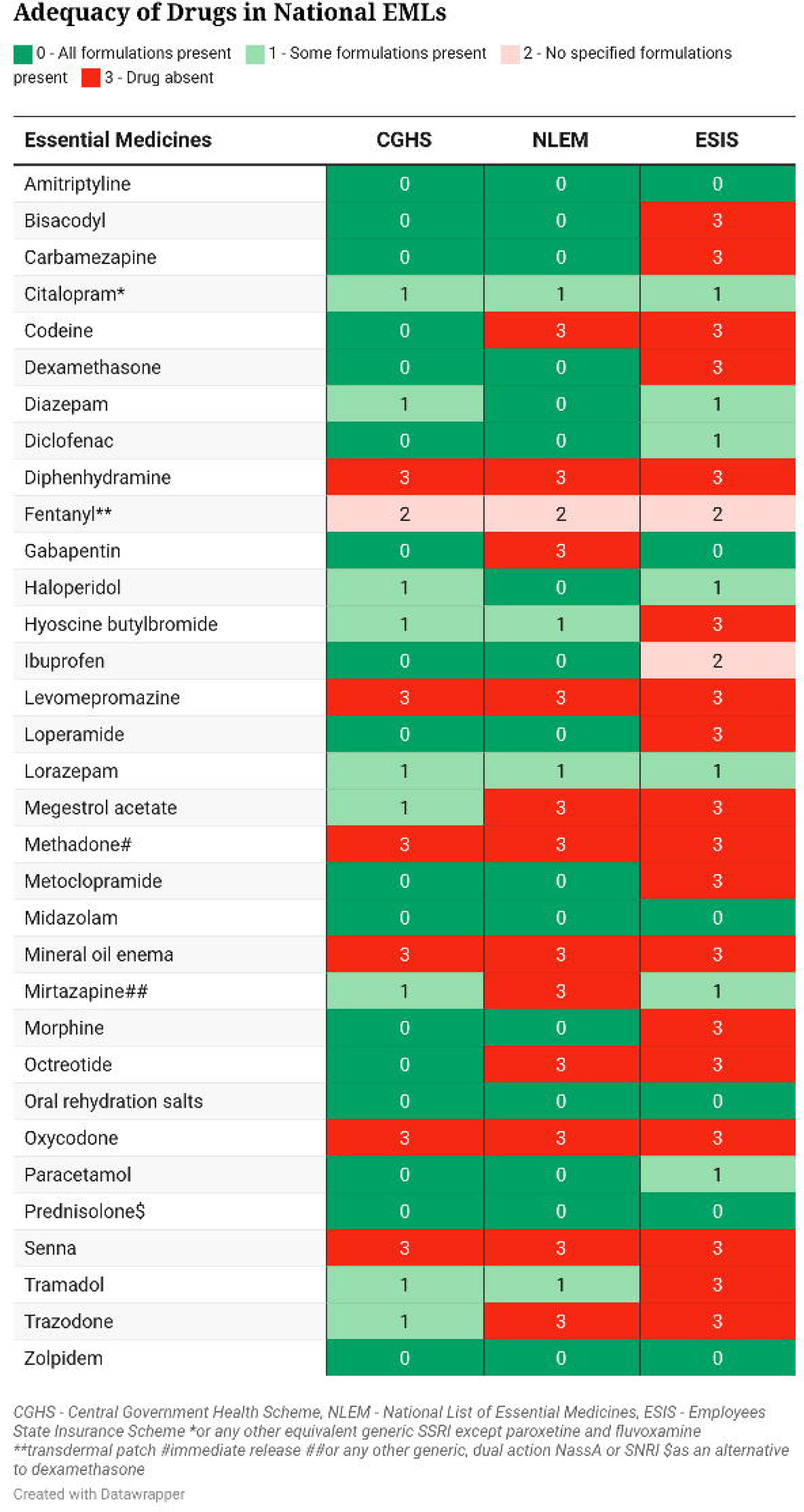
Adequacy of Indian national EMLs compared with the IAHPC EML for palliative care.

Among states and UTs, Delhi’s EML was the closest to the IAHPC EML and included all formulations required for 17 (52%) drugs. Eight (24%) drugs had some formulations present, one drug had no recommended formulation present, and seven (21%) drugs were absent. Nagaland’s EML included all formulations required for 3 (9%) drugs. Nine (27%) drugs had some formulations present, and 21 (64%) drugs were absent. The adequacy analysis of all EMLs is presented in **Figure 2** and **Table 3**. The presence of viable alternatives to absent drugs was variable across the EMLs, ranging from 29% of absent drugs in the Nagaland EML to 100% of absent drugs in CGHS, Delhi, and Uttarakhand EMLs.

**Figure 2:**
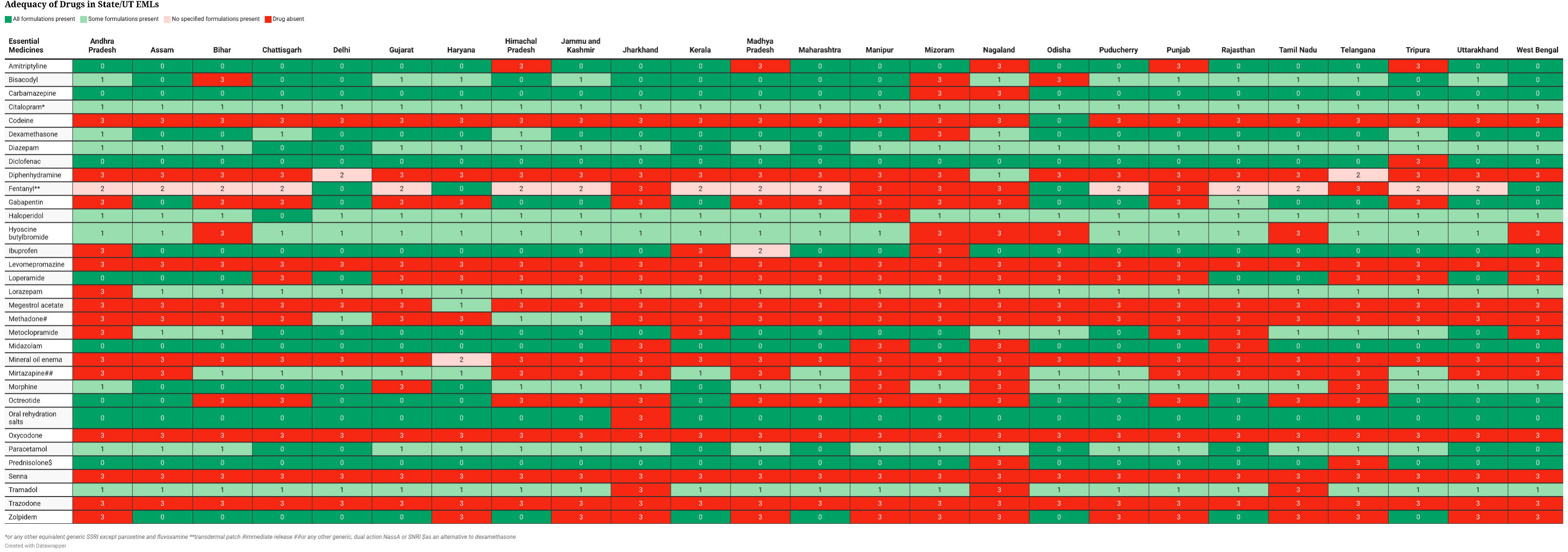
Adequacy of state/UT EMLs compared with the IAHPC EML for palliative care.

**Table 3:** Drug availability (%) compared with IAHPC recommendations.

| S. No. | EML | All formulations present | Some formulations present | No recommended formulation present | Drug absent |
| --- | --- | --- | --- | --- | --- |
|  | **National** |  |  |  |  |
| 1 | CGHS | 48 | 30 | 3 | 19 |
| 2 | NLEM | 46 | 15 | 3 | 36 |
| 3 | ESIS | 18 | 21 | 6 | 55 |
|  | **States and UTs** |  |  |  |  |
| 1 | Andhra Pradesh | 24 | 28 | 3 | 45 |
| 2 | Assam | 40 | 27 | 3 | 30 |
| 3 | Bihar | 30 | 27 | 3 | 40 |
| 4 | Chhattisgarh | 40 | 21 | 3 | 36 |
| 5 | Delhi | 52 | 24 | 3 | 21 |
| 6 | Gujarat | 33 | 27 | 3 | 37 |
| 7 | Haryana | 33 | 34 | 3 | 30 |
| 8 | Himachal Pradesh | 30 | 30 | 3 | 37 |
| 9 | Jammu and Kashmir | 30 | 30 | 3 | 37 |
| 10 | Jharkhand | 24 | 21 | 0 | 55 |
| 11 | Kerala | 39 | 22 | 3 | 36 |
| 12 | Madhya Pradesh | 24 | 24 | 7 | 45 |
| 13 | Maharashtra | 39 | 22 | 3 | 36 |
| 14 | Manipur | 27 | 18 | 0 | 55 |
| 15 | Mizoram | 18 | 21 | 0 | 61 |
| 16 | Nagaland | 9 | 27 | 0 | 64 |
| 17 | Odisha | 42 | 25 | 0 | 33 |
| 18 | Puducherry | 33 | 31 | 3 | 33 |
| 19 | Punjab | 21 | 27 | 0 | 52 |
| 20 | Rajasthan | 33 | 28 | 3 | 36 |
| 21 | Tamil Nadu | 30 | 24 | 3 | 43 |
| 22 | Telangana | 27 | 27 | 3 | 43 |
| 23 | Tripura | 25 | 33 | 3 | 39 |
| 24 | Uttarakhand | 39 | 24 | 3 | 34 |
| 25 | West Bengal | 39 | 19 | 0 | 42 |

No Indian EML had all recommended formulations of morphine as oral solutions, tablets, and injectables. Nationally, the CGHS EML and NLEM included morphine injectables and tablets, while the ESIS EML did not include morphine in any form. Eight (32%) state/UT EMLs included injectables and tablets. Twelve (48%) included only injectable morphine and one (3%) included only tablets. Morphine was absent in four (16%) EMLs - Gujarat, Manipur, Nagaland, and Telangana.

### 3.2 Adequacy for symptom management

All of the 16 symptoms listed by IAHPC could be managed by at least one drug present in two (67%) national lists - CGHS and NLEM and 18 (72%) state/UT EMLs. The analysis for symptom management in EMLs is presented in **Figure 3**. Among the inadequately managed symptoms, constipation was not addressed by the EMLs of ESIC, Bihar, and Mizoram. Moderate to severe pain management was insufficient in the EMLs of Manipur, Nagaland, and Telangana. Manipur’s EML did not manage terminal restlessness effectively, while Bihar’s EML did not manage both terminal respiratory congestion and visceral pain. Dyspnea was inadequately managed in the EMLs of ESIC, Gujarat, Manipur, Nagaland, and Telangana. Jharkhand’s EML was the only one that left diarrhoea unmanaged. However, depression, neuropathic pain, anorexia, nausea, vomiting, anxiety, mild to moderate pain, delirium, and insomnia were adequately managed by all state and national EMLs.

**Figure 3:**
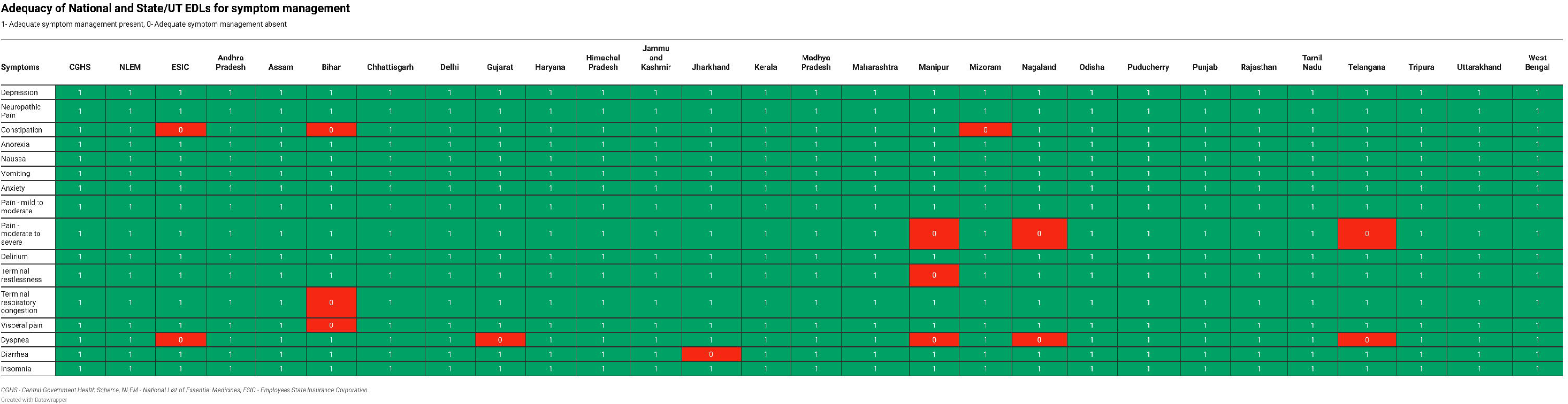
Adequacy of Indian EMLs for managing palliative care symptoms listed by IAHPC.

## 4. DISCUSSION

In our study, we report that India’s national and state EMLs did not completely align with the recommendations of IAHPC. The Delhi EML was the closest to IAHPC recommendations, followed by the NLEM. Some drugs recommended by the IAHPC such as levomepromazine, senna, trazodone, and oxycodone were absent from all state and national lists, except trazodone, which was present in the NLEM. Others such as codeine, megestrol, and mineral oil enema were present in only one state list each, and only the NLEM included codeine and megestrol. While EMLs include fentanyl in the injectable form, the commonly prescribed formulation for pain management - transdermal patch, was included by only one state (Haryana). The experts in the study felt that morphine was a more cost-effective and efficient substitute for transdermal fentanyl in the Indian setting due to the difficulty of titration and the high cost of the latter, which restricts access to it. Additionally, even though all EMLs, barring one, contained ORS for the management of diarrhoea, Indian EMLs were found to be inadequate to treat intractable diarrhoea, since the presence of loperamide is integral for non-infective intractable diarrhoea management. (19,20) Dyspnea, constipation, and moderate to severe pain are other symptoms poorly managed by Indian EMLs. States such as Bihar, Telangana, and the north-eastern states of Manipur, Nagaland, and Mizoram lag behind the rest of the country in the inclusion of recommended drugs, as well as symptom management, even after including viable alternatives present in the EMLs.

Essential drugs cater to any population’s priority needs, availability, and affordability. Given the rising number of patients needing palliative care, essential drugs must be made available and accessible to them. EMLs are instrumental in improving access to essential drugs through the public health system and reducing the financial burden on patients and caregivers for conditions that are relatively common in the community. (21,22) Globally, the implementation of EMLs has improved access to treatment, with improved drug availability, better prescription practices, and reduced costs. The implementation of the National Essential Medicines Policy in China led to better prescription practices and a decline in average prescription costs. Another study looking at the impact of the essential drugs programme in peripheral health units in Yemen showed similar results, with improved availability and more rational use of drugs. (23,24)

While we used the IAHPC EML, published in 2007, as the benchmark against which all other EMLs were compared, we noticed that certain updates based on recent evidence need to be incorporated. For example, in 2010, Fosbøl et al. reported an increased risk of cardiovascular events in patients receiving treatment with diclofenac which has been mentioned in the IAHPC list for the management of mild to moderate pain. (25) Additionally, we propose a further classification of nausea and vomiting into that caused by gastroparesis, radiotherapy, and chemotherapy for simplification of prescription, as their treatments widely differ. (26)

The increasing advocacy for fentanyl transdermal patches, touted for their potency in pain management, is raising concerns, particularly in low- and middle-income countries (LMICs) like India. While a 100-microgram dose of fentanyl is equivalent to 10 grams of morphine,(27) in India fentanyl costs 12 times as much as morphine per day. The high cost of fentanyl, coupled with its limited accessibility, makes it impractical for widespread use in LMICs. (27,28) Indian EMLs did not include all the recommended formulations of morphine, a practical alternative to fentanyl. Morphine’s importance is highlighted by the fact that an objective of the National Program for Palliative Care (NPPC) is to increase its availability. Morphine usage is also used as an indicator to assess the coverage of palliative care services in the NPNCD. (29) Emphasis should be placed on ensuring that all formulations of morphine are available and affordable. Despite the 2014 amendment to the NDPS Act, which reduced the number of required licences for opioid possession from six to one, complex regulatory procedures and a lack of prescription awareness among physicians continue to hinder patients’ access to essential pain management medications like morphine. In 2014, India’s total morphine consumption was a mere 278 kg. Considering that a patient with terminal cancer requires 75 mg of morphine per day for approximately 90 days, this amount is only sufficient to adequately treat 40,000 patients. (30,31) Addressing these barriers is crucial for improving pain management in countries like India, where the need for affordable and effective pain relief is paramount. Simplifying regulatory processes and enhancing physician education on opioid prescriptions are key steps toward ensuring that patients receive the pain management they need.

The Sustainable Development Goal 3.8 mentions access to quality essential healthcare services and access to safe, effective, quality, and essential medicines for the achievement of Universal Health Coverage (UHC). We noted the presence of different national EMLs, meant effectively for different sections of the population, with the CGHS EML meant for government employees performing better than the ESIS EML meant for other workers. This highlights redundancy in the system and a potential source for inequitable healthcare delivery. Additionally, the EMLs devised by individual states should ideally include all the drugs present in the national list, with the addition of drugs considered necessary based on the local epidemiology of diseases. Establishing a single national EML, aligned with global standards is integral to achieving UHC, especially for palliative care provision. By implementing EMLs tailored to include drugs required to deliver palliative care, healthcare systems can effectively address the diverse needs of patients while promoting equitable and cost-effective healthcare delivery. (22)

India launched NPPC in 2012 to make high-quality palliative care accessible across all levels of health care. (29) This included making drugs for pain relief and other symptoms available at the primary healthcare level. (32) Thus, it is imperative that the benefits of drug inclusion in the EML are not restricted to tertiary care setups, and that drugs are made available in primary and secondary care centres as well. (29) While national and state EMLs do not conform to the IAHPC recommendations, they contain alternatives for the management of symptoms commonly encountered in palliative care. However, some states, such as Bihar, Gujarat, Jharkhand, Mizoram, Manipur, Nagaland, and Telangana, lack these alternatives, rendering them inadequate in managing some of these symptoms. It is crucial to incorporate essential medications from the IAHPC list or adopt alternatives to address these gaps. For instance, adding morphine to Nagaland’s EML would improve the management of both moderate to severe pain and dyspnea. Furthermore, Jharkhand should include ORS in its EML for the management of diarrhoea, as it is the only EML currently lacking this essential treatment.

Our findings highlight that there is a need for an EML tailored to palliative care in India and similarly in other countries, with drugs thoughtfully included as per existing procurement, storage, and distribution constraints. EMLs specific for settings with different levels of available resources will help in guiding countries that currently do not have an EML or are looking to update existing EMLs.

### Strengths and Limitations

Our study is the first to analyse the presence of specific medications essential for palliative care across the diverse EMLs of India. This contributes significantly to evaluating their readiness to manage the prevalent symptoms encountered by patients receiving palliative care. We were also able to delve deeper into analysing the management of common symptoms experienced by patients receiving palliative care as in cases where the drugs recommended by the IAHPC were unavailable, alternative options were explored.

Our study has a few limitations. First, while we identified the inclusion of medications for palliative care across multiple EMLs, we lacked the resources to assess their availability at the grassroots level and within hospitals. A comprehensive evaluation, combining our study with grassroots-level investigations, would provide a more accurate measure of access to essential drugs. Second, we did not compare the available drug doses in EMLs with the recommendations of IAHPC as the required dose can be attained by modifying the quantity consumed of the available drug. Lastly, the costs of the alternate drugs listed by experts were not compared. However, it was a criterion that the experts considered for deciding the alternatives.

## 5. CONCLUSION

Indian EMLs are not entirely in line with the IAHPC recommendations for essential palliative care drugs. However, they contain a range of drugs adequate to treat most symptoms requiring palliation. Considering the importance of morphine, both in palliative care symptom management and monitoring of palliative care-related national programs in India, the national and state/UT EMLs should be updated to incorporate oral and injectable formulations of morphine. There is a need to update the IAHPC list using recent evidence, and there is also a need to design a list based on different levels of available resources to guide countries in formulating their EMLs for palliative care service delivery.

## 6. AUTHORSHIP

Corresponding author: PS

Joint authorship: DA and DS contributed equally to this paper.

Conceptualization: PS; Reviewed study proposal: PS, SZ, MRR, AG; Data extraction: DA, DS; Data analysis: DA, DS; Scientific advisors: MRR, AG, SZ; Writing of the original draft: DA, DS; Review and editing of the final draft: All authors; Project supervision: PS, SZ.

## 8. DATA SHARING

The data are available upon reasonable request that can be made to Dr Parth Sharma (Email ID)

## 9. ETHICAL STATEMENT

The study did not involve any human or animal subjects and therefore did not require any ethical clearance.

## Data Availability

Data are available upon reasonable request

## REFERENCES

1. WHO. Palliative care [Internet]. Palliative care. [cited 2024 Jan 17]. Available from: https://www.who.int/news-room/fact-sheets/detail/palliative-care

2. Maeda, et al I. Changes in Relatives’ Perspectives on Quality of Death, Quality of Care, Pain Relief, and Caregiving Burden Before and After a Region-Based Palliative Care Intervention. J Pain Symptom Manage. 2016 Sep 21;52(5).

3. Noble, et al H. Clinician views of patient decisional conflict when deciding between dialysis and conservative management: Qualitative findings from the PAlliative Care in chronic Kidney diSease (PACKS) study. sage journals. 2017 Apr 18;

4. Lalani N, Cai Y. Palliative care for rural growth and wellbeing: identifying perceived barriers and facilitators in access to palliative care in rural Indiana, USA. BMC Palliat Care. 2022 Feb 19;21(1):25.

5. Bag S, Mohanty S, Deep N, Salins N, Bag S. Palliative and End of Life Care in India - Current Scenario and the Way Forward. J Assoc Physicians India. 2020 Nov;68(11):61–5.

6. Patil S, Sharma P, Arora A, Zadey S. Unmet need for cancer palliative care in India. BMJ Support Palliat Care. 2024 Jun 8;

7. WHO Model Lists of Essential Medicines [Internet]. [cited 2024 Jun 7]. Available from: https://www.who.int/groups/expert-committee-on-selection-and-use-of-essential-medicines/essential-medicines-lists

8. Kar SS, Pradhan HS, Mohanta GP. Concept of essential medicines and rational use in public health. Indian J Community Med. 2010 Jan;35(1):10–3.

9. Constitution of India- Seventh Schedule.

10. IAHPC Essential Medicines for Palliative Care - International Association for Hospice & Palliative Care [Internet]. [cited 2024 Feb 17]. Available from: https://hospicecare.com/what-we-do/projects/palliative-care-essentials/iahpc-essential-medicines-for-palliative-care/

11. Vision and Mission - International Association for Hospice & Palliative Care [Internet]. [cited 2024 May 28]. Available from: https://hospicecare.com/about-iahpc/who-we-are/vision-and-mission/

12. Ministry of Health and Family Welfare, Government of India. National List of Essential Medicines [Internet]. 2022 [cited 2024 Jun 13]. Available from: https://main.mohfw.gov.in/sites/default/files/Notification%20and%20Report%20on%20National%20List%20of%20Essential%20Medicines%2C%202022.pdf

13. Employees State Insurance Corporation (ESIC). Employee’s State Insurance Corporation: Medical Rate Contract [Internet]. 2010 [cited 2024 Jun 13]. Available from: https://www.esic.gov.in/Tender/rc135data.pdf

14. Directorate General of Health Services. CGHS list of Life saving drugs [Internet]. [cited 2024 Jun 13]. Available from: https://cghs.nic.in/REVISED%20LIST%20OF%20LIFESAVING%20DRUGS%20OF%20CGHS%20MSD%20DELHI.pdf

15. Liliana De Lima et al. Ensuring Palliative Medicine Availability: The Development of the IAHPC List of Essential Medicines for Palliative Care. Jornal of Pain and Symptom Management. 2007 May;33(5).

16. Dalglish SL, Khalid H, McMahon SA. Document analysis in health policy research: the READ approach. Health Policy Plan. 2021 Feb 16;35(10):1424–31.

17. Iwona Zaporowska-Stachowiak et al. Haloperidol in palliative care: Indications and risks. Biomed Pharmacother. 2020 Dec;Volume 132.

18. Ministry of Health and Family Welfare, Government of India. Operational guidelines National Programme for Prevention and Control of Non-Communicable Diseases [Internet]. [cited 2024 May 28]. Available from: https://media.licdn.com/dms/document/media/D4D1FAQFUE2ll_2hsmg/feedshare-document-pdf-analyzed/0/1684375232945?e=1717632000&v=beta&t=Zt6cZJnsCoNPyOrsopQ9-tswx99fQXtsJDDUgN1lA

19. Maroun JA, Anthony LB, Blais N, Burkes R, Dowden SD, Dranitsaris G, et al. Prevention and management of chemotherapy-induced diarrhea in patients with colorectal cancer: a consensus statement by the Canadian Working Group on Chemotherapy-Induced Diarrhea. Curr Oncol. 2007 Feb;14(1):13–20.

20. Clay PG, Crutchley RD. Noninfectious diarrhea in HIV seropositive individuals: a review of prevalence rates, etiology, and management in the era of combination antiretroviral therapy. Infect Dis Ther. 2014 Dec;3(2):103–22.

21. Jhaj R, Banerjee A, Kshirsagar NA, Sadasivam B, Chandy SJ, Bright HR, et al. Use of drugs not listed in the National List of Essential Medicines: Findings from a prescription analysis by the Indian Council of Medical Research-Rational Use of Medicines Centres Network in tertiary care hospitals across India. Indian J Pharmacol. 2022;54(6):407–16.

22. Wirtz VJ, Hogerzeil HV, Gray AL, Bigdeli M, de Joncheere CP, Ewen MA, et al. Essential medicines for universal health coverage. Lancet. 2017 Jan 28;389(10067):403–76.

23. Chao J, Gu J, Zhang H, Chen H, Wu Z. The impact of the national essential medicines policy on rational drug use in primary care institutions in jiangsu province of china. Iran J Public Health. 2018 Jan;47(1):24–32.

24. Hogerzeil HV, Walker GJ, Sallami AO, Fernando G. Impact of an essential drugs programme on availability and rational use of drugs. Lancet. 1989 Jan 21;1(8630):141–2.

25. Fosbøl EL, Folke F, Jacobsen S, Rasmussen JN, Sørensen R, Schramm TK, et al. Cause-specific cardiovascular risk associated with nonsteroidal antiinflammatory drugs among healthy individuals. Circ Cardiovasc Qual Outcomes. 2010 Jul;3(4):395–405.

26. Sik Kim Ang et al. Nausea and Vomiting in Advanced Cancer. Am J Hosp Palliat Care. 2010 Mar 2;

27. Ramos-Matos CF, Bistas KG, Lopez-Ojeda W. Fentanyl. StatPearls. Treasure Island (FL): StatPearls Publishing; 2024.

28. Deirdre M Neighbors et al. Economic Evaluation of the Fentanyl Transdermal System for the Treatment of Chronic Moderate to Severe Pain. J Pain Symptom Manage. 2001 Feb;VOLUME 21(ISSUE 2):P129-143.

29. Directorate General Of Health Services [Internet]. [cited 2024 May 8]. Available from: https://dghs.gov.in/content/1351_3_NationalProgramforPalliativeCare.aspx

30. Jacob A, Mathew A. End-of-Life Care and Opioid Use in India: Challenges and Opportunities. JCO Global Oncology. 2017 Jan 25;

31. India: The principle of balance to make opioids accessible for palliative care [Internet]. [cited 2024 Jul 7]. Available from: https://www.unodc.org/southasia/frontpage/2011/april/interview-m-r-rajagopal-access-to-o pioids-for-palliative-care.html

32. Ministry of Health and Family Welfare- Government of India. Operational Guidelines: Palliative care at Health and Wellness Centers [Internet]. [cited 2024 May 29]. Available from: https://nhsrcindia.org/sites/default/files/2021-06/Operational%20Guidelines%20for%20Pa lliative%20Care%20at%20HWC.pdf

