## Supplementary table 1 for "An evaluation of the adequacy of Indian national and state Essential Medicines Lists (EMLs) for palliative care medical needs - a comparative analysis"

**Supplementary Table 1: National and State Essential Medication Lists.**

| S. No. | National List | Year | Authorizing body | Reference |
|---|---|---|---|---|
| 1 | Central Government Health Scheme (CGHS) | Unknown | Directorate General of Health Services | https://cghs.nic.in/REVISED%20LIST%20OF%20LIFESAVING%20DRUGS%20OF%20CGHS%20MSD%20DELHI.pdf |
| 2 | Employees State Insurance Scheme (ESIS) | 2010 | Employees State Insurance Corporation (ESIC) | https://www.esic.gov.in/Tender/rc135data.pdf |
| 3 | National List of Essential Medicines (NLEM) | 2022 | Ministry of Health and Family Welfare, Government of India | https://main.mohfw.gov.in/sites/default/files/Notification%20and%20Report%20on%20National%20List%20of%20Essential%20Medicines%2C%202022.pdf |
| **S. No.** | **State/Union Territory** | **Year** | **Authorizing body** | **Reference** |
| 1 | Andhra Pradesh | 2013 | Health and Family Welfare Department, Government of Andhra Pradesh | https://cfw.ap.nic.in/pdf/Employees%20Data/Free%20D&D/2013HMF_MS204.pdf |
| 2 | Assam | 2023 | Medical Education and Research Department, Government of Assam | https://amscl.assam.gov.in/sites/default/files/public_utility/Notification%20No.MER_.424912-72%20-%20Essential%20Drug%20List%20%28EDL%29%20and%20Specialty%20Drug%20List%20%28SDL%29%202023-24_compressed.pdf |
| 3 | Bihar | 2022 | Health Department, Government of Bihar | https://shs.bihar.gov.in/SHS/Drug_Equipments/Sankalp_1729_12_Dt-08-12-2022-EDL-2022.pdf |

| | | | | |
|---|---|---|---|---|
| 4 | Chhattisgarh | 2016 | Department of Health and Family Welfare Department, Government of Chhattisgarh | https://cgmsc.gov.in/Upload/EDL%202016.pdf |
| 5 | Delhi | 2022 | Directorate of Health Services, Government of Delhi | https://dgehs.delhi.gov.in/dghs/edl |
| 6 | Gujarat | 2022 | Government of Gujarat | https://gmscl.gujarat.gov.in/essential-drug-list.htm |
| 7 | Haryana | 2013 | Government of Haryana | https://hmscl.org.in/Edl.html |
| 8 | Himachal Pradesh | 2020 | Health and Family Welfare Department, Government of Himachal Pradesh | https://himachal.nic.in/WriteReadData/l892s/19_l892s/1593418077.pdf |
| 9 | Jammu and Kashmir | 2022 | Health and Medical Education Department, Government of Jammu and Kashmir | https://www.jkmsclbusiness.com/pdf/jkmscledl22jan22.pdf |
| 10 | Jharkhand | | Government of Jharkhand | https://jmhidpcl.jharkhand.gov.in/uploads/Essential-Drug-List-Jharkhand.pdf |
| 11 | Kerala | 2020 | Government of Kerala | http://kmscl.kerala.gov.in/KMSCL/uploads/announcements/DRUG LIST_2020-21_-_WEBSITE.pdf |
| 12 | Madhya Pradesh | 2020 | Directorate of Health Services, Government of Madhya Pradesh | https://www.sda.mp.gov.in/mphealth/en/drug-procurement/essential-drug-listpolicy |

| | | | | |
|---|---|---|---|---|
| 13 | Maharashtra | 2022 | Commissionerate of Health Services, Government of Maharashtra | https://nrhm.maharashtra.gov.in/EDL.pdf |
| 14 | Manipur | 2012 | State Health Society, Manipur | https://nrhmmanipur.org/wp-content/uploads/2014/04/ANNEXURE-medicine-list-for-CHC-PHC-PHSC-UHC.pdf |
| 15 | Mizoram | 2023 | Mizoram Health and Family Welfare Department | https://health.mizoram.gov.in/uploads/attachments/2023/07/2d717add94f748f20debac8321ab6f06/essential-drugs.pdf |
| 16 | Nagaland | 2018 | Health and Family Welfare Department, Government of Nagaland | https://www.nhmnagaland.in/Notification_file_path/FACILITY%20TYPE%20ESSENTIAL%20DRUG%20LIST.pdf |
| 17 | Odisha | 2020 | Odisha Health and Family Welfare Department | https://osmcl.nic.in/sites/default/files/Essential%20Drug%20List%202020.pdf |
| 18 | Puducherry | 2023 | Department of Health and Family Welfare Services, Government of Puducherry | https://health.py.gov.in/revised-essential-drug-list-edl |
| 19 | Punjab | 2018 | Government of Punjab | https://phsc.punjab.gov.in/sites/default/files/2019-08/520183121639ListofEDL%26JSSKitems2018.xls |
| 20 | Rajasthan | Unknown | Government of Rajasthan | http://rmsc.health.rajasthan.gov.in/content/dam/doitassets/Medical-and-Health-Portal/rajasthan-medical-corporation/pdf/EDL/EDL%20final%20list%203.3.2020.pdf |

| | | | | |
|---|---|---|---|---|
| 21 | Tamil Nadu | 2022 | Government of Tamil Nadu | https://tnmsc.tn.gov.in/user_pages/drugtender.php?drugcat=T21083 |
| 22 | Telangana | Unknown | Department of Health, Medical and Family Welfare, Government of Telangana | http://tsmsidc.telangana.gov.in/open_record_view.php?ID=178 |
| 23 | Tripura | 2017 | Health and Family Welfare Department, Government of Tripura | https://drive.google.com/file/d/10vH9hytgU6OM2o5gYhiiupJXGaL4x3Rs/view?usp=sharing |
| 24 | Uttarakhand | 2015 | Department of Medical Health and Family Welfare, Government of Uttarakhand | https://nhm.uk.gov.in/upload/tenders/Tender-11.pdf |
| 25 | West Bengal | 2022 | Department of Health and Family Welfare, Government of West Bengal | https://www.wbhealth.gov.in/uploaded_files/go/EDL_507.pdf |
